# Rethinking Transfer Learning for Medical Image Classification

**DOI:** 10.1101/2022.11.26.22282782

**Authors:** Le Peng, Hengyue Liang, Gaoxiang Luo, Taihui Li, Ju Sun

**Author notes:** https://lepeng.org. https://hengyuel.github.io. https://gaoxiangluo.github.io. https://taihui.github.io. https://sunju.org.

## Abstract

Transfer learning (TL) from pretrained deep models is a standard practice in modern medical image classification (MIC). However, what levels of features to be reused are problem-dependent, and uniformly finetuning all layers of pretrained models may be suboptimal. This insight has partly motivated the recent *differential* TL strategies, such as TransFusion (TF) and layer-wise finetuning (LWFT), which treat the layers in the pretrained models differentially. In this paper, we add one more strategy into this family, called *TruncatedTL*, which reuses and finetunes appropriate bottom layers and directly discards the remaining layers. This yields not only superior MIC performance but also compact models for efficient inference, compared to other differential TL methods. Our code is available at: https://github.com/sun-umn/TTL.

## 1 Introduction

Transfer learning (TL) is a common practice for medical image classification (MIC), especially when training data are limited. In typical TL pipelines for MIC, deep convolutional neural networks (DCNNs) pretrained on large-scale *source tasks* (e.g., object recognition on ImageNet [6]) are finetuned as backbone models for *target MIC tasks*; see, e.g., [1, 9, 14, 27, 31, 39], for examples of prior successes.

The key to TL is feature reuse from the source to the target tasks, which leads to practical benefits such as fast convergence in training, and good test performance even if the target data are scarce [43]. Pretrained DCNNs extract increasingly more abstract visual features from bottom to top layers: from low-level corners and textures, to mid-level blobs and parts, and finally to high-level shapes and patterns [43, 44]. While shapes and patterns are crucial for recognizing and segmenting generic visual objects (see Fig. 1 (left)), they are not necessarily the defining features for diseases: diseases can often take the form of abnormal textures and blobs, which correspond to low-to mid-level features (see Fig. 1 (right)). *So intuitively for MIC, we may only need to finetune a reasonable number of the bottom layers commensurate with the levels of features needed, and ignore the top layers*. However, standard TL practice for MIC retains all layers, and uses them as fixed feature extractors or finetunes them uniformly.

**Figure 1.**
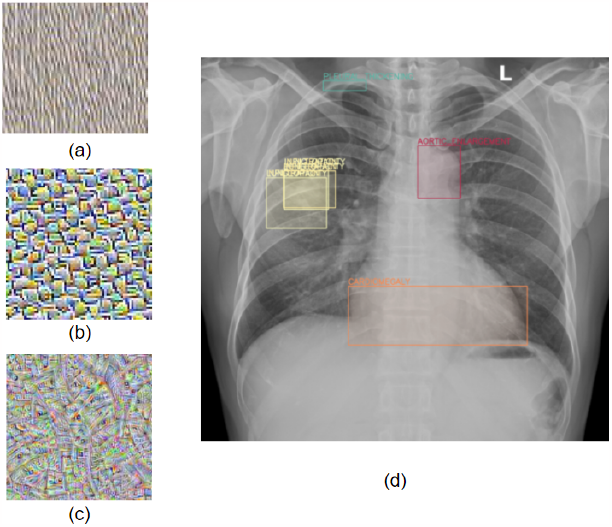
(left) The feature hierarchy learned by typical DCNNs, see Appendix E for details; (right) Examples of diseases in a chest x-ray [21].

Raghu et al. [26], Tajbakhsh et al. [34] depart from the uniform TL approach and propose TL methods that treat top and bottom layers differently. Prioritizing high-level features and the classifier, Tajbakhsh et al. [34] proposes *layer-wise finetuning* (LWFT) that finetunes an appropriate number of top layers and freezes the remaining bottom layers. In comparison, to improve training speed while preserving performance, Raghu et al. [26] proposes *Trans-Fusion* (TF) that finetunes bottom layers but retrains a coarsened version of top layers from scratch.

Neither of the above *differential* TL strategies clearly address the conundrum of why top layers are needed when only the features in bottom layers are to be reused. To bridge the gap, in this paper, we propose a novel, perhaps radical, TL strategy: remove top layers after an appropriate cutoff point, and finetune the truncated model left, dubbed *TruncatedTL* (TTL)—this is entirely consistent with our intuition about the feature hierarchy. Our main contributions include: 1) **confirming the deficiency of full TL**. By experimenting with full and differential TL strategies—including our TTL—on three MIC tasks and three popularly used DCNN models for MIC, we find that full TL (FTL) is almost always suboptimal in terms of classification performance, confirming the observation in Tajbakhsh et al. [34]. 2) **proposing TruncatedTL (TTL) that leads to effective and compact models**. Our TTL outperforms other differential TL methods, while the resulting models are always smaller, sometimes substantially so. This leads to reduced computation and fast prediction during inference, and can be particularly valuable when dealing with 3D medical data such as CT and MRI images. 3) **quantifying feature transferability in TL for MIC**. We use singular vector canonical correlation analysis (SVCCA) [24] to analyze feature transferability and confirm the importance of low- and mid-level features for MIC. The quantitative analysis also provides insights on how to choose high-quality truncation points to further optimize the model performance and efficiency of TTL.

## 2 Related Work

**Deep TL** TL by reusing and finetuning visual features learned in DCNNs entered computer vision (CV) once DCNNs became the cornerstone of state-of-the-art (SOTA) object recognition models back to 2012. For example, Donahue et al. [7], Sharif Razavian et al. [29] propose using part of or full pretrained DCNNs as feature extractors for generic visual recognition. Yosinski et al. [43] studies the hierarchy of DCNN-based visual features, characterizes their transferability, and proposes finetuning pretrained features to boost performance on target tasks. Moreover, Erhan et al. [8], Zeiler and Fergus [44] propose techniques to visualize visual features and their hierarchy. This popular line of TL techniques is among the broad family of TL methods for knowledge transfer from source to target tasks based on deep networks [35] and other learning models [45], and is the focus of this paper.

### General TL stategies

While pretrained DCNNs can be either used as fixed feature extractors, or partially or fully finetuned on the target data, Yosinski et al. [43] argues that bottom layers learn generic features while top layers learn task-specific features, leading to folklore guidelines on how to choose appropriate TL strategies in various scenarios, as summarized in Fig. 2.

**Figure 2.**
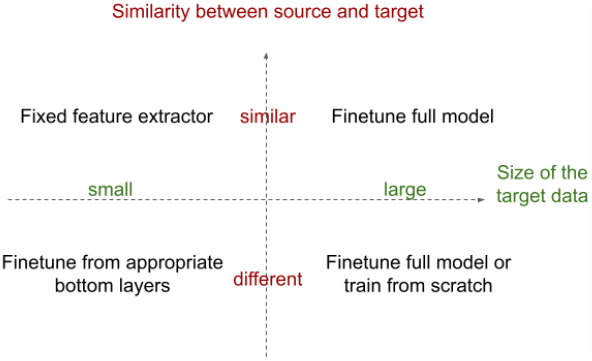
TL strategies and their usage scenarios.

Our intuition about the feature hierarchy is slightly different: bottom layers learn low-level features that are spatially localized, and top layers learn high-level features that are spatially extensive (see Fig. 1). Although the two hierarchies may be aligned for most cases, they are distinguished by whether spatial scales are considered: task-specific features may be spatially localized, e.g., texture features to classify skin lesions [36], and general features may be spatially extensive, e.g., generic brain silhouettes in brain MRI images. Moreover, the spatial-scale hierarchy is built into DCNNs by design [3]. So we argue that the intuition about the low-high spatial feature hierarchy is more pertinent. We note that none of the popular strategies as summarized in Fig. 2 modify the pretrained DCNN models (except for the final multi-layer perceptron, MLP, classifiers)—in contrast to TF and our TTL.

### Differential TL

Early work on TL for MIC [1, 11, 14, 27, 31, 33, 39] and segmentation [9, 16, 17, 28, 38] parallels the relevant developments in CV, and mostly uses DCNNs pretrained on CV tasks as feature extractors or initializers (i.e., for finetuning). In fact, these two strategies remain dominant according to the very recent survey [18] on TL for MIC, which reviews around 120 relevant papers. But the former seems inappropriate as medical data are disparate from natural images dealt with in CV. From the bottom panel of Fig. 2, finetuning at least part of the DCNNs is probably more competitive even if the target data are limited. In this line, LWFT [34] finetunes top layers and freeze bottom layers, and incrementally allows finetuning more layers during model selection—this is recommended in Kim et al. [18] as a practical TL strategy for MIC that strikes a balance between training efficiency and performance. Similarly, TF [26] coarsens top layers which are then trained from scratch, and finetunes bottom layers from pretrained weights. Both LWFT and TF take inspiration from the general-specific feature hierarchy. In contrast, motivated by the low-high spatial feature hierarchy, our novel TTL method removes the top layers entirely and directly finetunes the truncated models. Our experiments in Section 4 confirm that TTL surpasses LWFT and TF with improved performance and reduced inference cost.

### Compact models for MIC

Both TF and our TTL lead to reduced models that can boost the inference efficiency, the first of its kind in TL for MIC, although the TF paper [26] does not stress this point. Compact models have been designed for specific MIC tasks, e.g., Huang et al. [13], Raghu et al. [26], but our evaluation on the task of Huang et al. [13] in Section 4 suggests that the differential TL strategies based on generic pretrained models, particularly our TTL, can outperform TL based on handcrafted models. Moreover, the growing set of methods for model quantization and compression [10, 19, 23, 41] are equally applicable to both the original models and the reduced models.

## 3 Efficient transfer learning for MIC

Let *𝒳 ×𝒴* denote any input-output (or feature-label) product space, and *D*_*𝒳×𝒴*_ a distribution on *𝒳 × 𝒴*. TL considers a source task 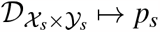, where *p*_*s*_ is a desired predictor, and a target task 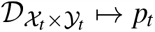. In typical TL, *𝒳*_*t*_ *× 𝒴*_*t*_ may be different from *𝒳*_*s*_ *× 𝒴*_*s*_, or at least 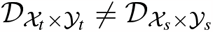 even if *𝒳*_*t*_ *× 𝒴*_*t*_ = *𝒳*_*s*_ *× 𝒴*_*s*_. The goal of TL is to transfer the knowledge from the source task 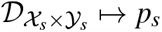 that is solved beforehand to the target task 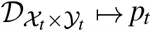 [22, 35].

In this paper, we restrict TL to reusing and finetuning pretrained DCNNs for MIC. In this context, the source predictor *p*_*s*_ = *h*_*s*_ ◯ *f*_*L*_ ◯ *···* ◯ *f*_1_ is pretrained on a large-scale source dataset 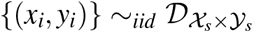. Here, the *f*_*i*_’s are *L* convolulational layers, and *h*_*s*_ is the final MLP classifier. To perform TL, *h*_*s*_ is replaced by a new MLP predictor *h*_*t*_ with prediction heads matching the target task (i.e., with the desired number of outputs) to form the new model *p*_*t*_ = *h*_*t*_ ◯ *f*_*L*_ ◯ *· · ·* ◯ *f*_1_. Two dominant approaches of TL for MIC are: 1) *fixed feature extraction*: freeze the pretrained weights of *f*_*L*_ ◯ *· · ·* ◯ *f*_1_, and optimize *h*_*t*_ from random initialization so that *p*_*t*_ fits the target data; 2) *full transfer learning (FTL)*: optimize all of *h*_*t*_ ◯ *f*_*L*_ ◯ *· · ·* ◯ *f*_1_, with *h*_*t*_ from random initialization whereas *f*_*L*_ ◯ *· · ·* ◯ *f*_1_ from their pretrained weights so that *p*_*t*_ fits the target data.

### 3.1 Prior differential TL approaches

The two differential TL methods for MIC, i.e., LWFT [34] and TF [25], differ from the dominant TL approaches in that they treat the top and bottom layers differently, as illustrated in Fig. 3 (i).

**Figure 3.**
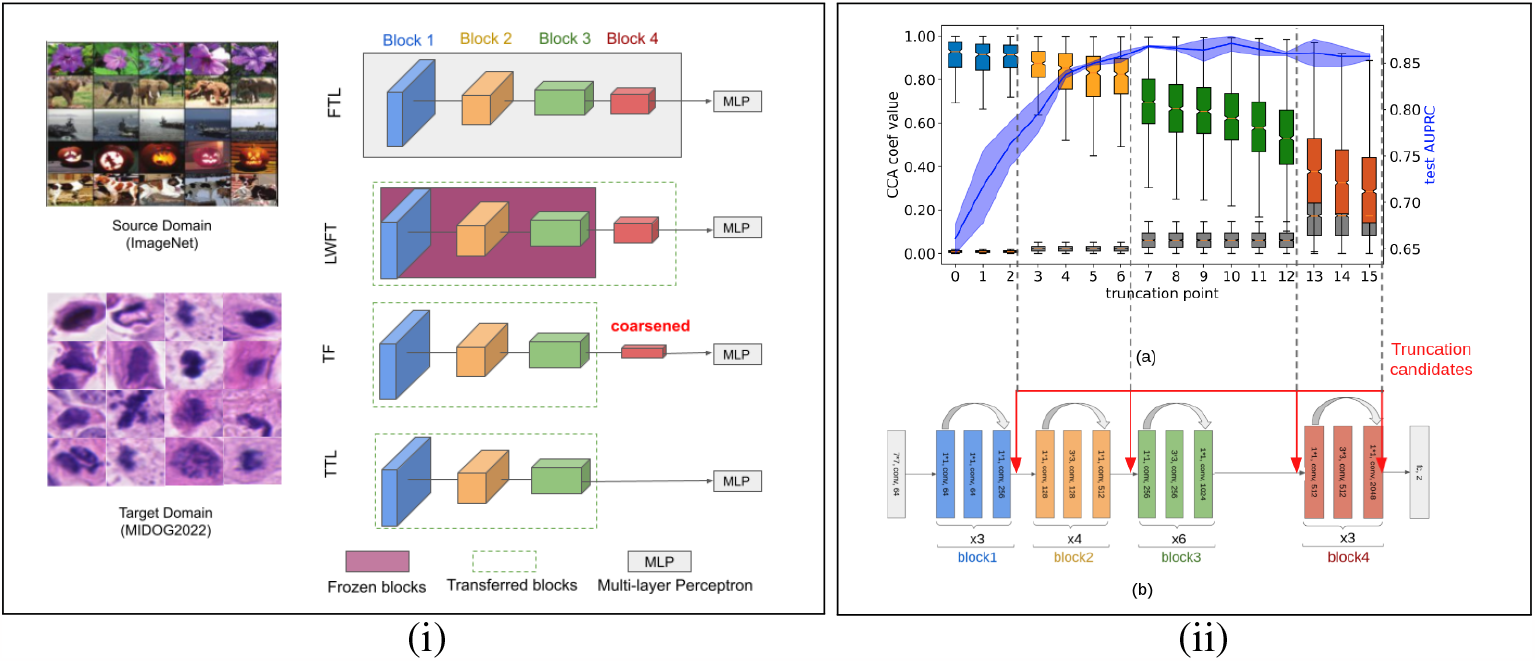
**(i)** Overview of typical TL setup, and the four TL methods that we focus on in this paper. **(ii)** Illustration of feature transferability and the performance of different levels of features on BIMCV. We take the ResNet50 model pretrained on ImageNet, and perform a full TL on BIMCV. We consider 17 natural truncation/cutoff points that do not cut through the skip connections.

### Layer-wise finetuning

LWFT does not distinguish MLP layers and convolutional layers. So slightly abusing our notation, assume that the pretrained DCNN is *f*_*N*_ ◯ *f*_*N−*1_ ◯ *· · ·* ◯ *f*_1_, where *N* is the total number of layers including both the MLP and convolutional layers. LWFT finetunes the top *k* layers *f*_*N*_ ◯ *f*_*N−*1_ ◯*···* ◯ *f*_*N−k*+1_, and freezes the bottom *N −k* layers *f*_*N−k*_ ◯ *· · ·* ◯ *f*_1_. The top layer *f*_*N*_ is finetuned with a base learning rate (LR) *η*, and the other *k −* 1 layers with a LR *η/*10. To find an appropriate *k*, Tajbakhsh et al. [34] proposes an in-cremental model selection procedure: start with *k* = 1 layer, and include one more layer into finetuning if the previous set of layers does not achieve the desired level of performance^1^. Although LWFT was originally proposed for AlexNet [20], it can be generalized to work with advanced DCNN models such as ResNets and DenseNets that have block structures.

### TransFusion

TF reuses bottom layers while slimming down top layers. Formally, for a cutoff index *k*, the pretrained model *p*_*s*_ ◯ *f*_*L*_ ◯ *· · ·* ◯ *f*_*k*+1_ ◯ *f*_*k*_ *· · ·* ◯ *f*_1_ is replaced by 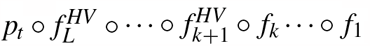, where HV means halving the number of channels in the designated layer. TF then trains the coarsened model on the target data with the first half *f*_*k*_ *· · ·* ◯ *f*_1_ initialized by the pretrained weights (i.e., finetuning) and 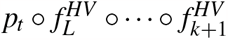 initialized by random weights (i.e., training from scratch). The cutoff point *k* is the key hyperparameter in TF. TF was originally proposed to boost the finetuning speed, but we find that it often also boosts the classification performance compared to FTL; see Section 4.

### 3.2 Our truncated TL approach

Our method is radically simple: for an appropriate cutoff index *k*, we take the *k* bottom layers *f*_*k*_ ◯ *· · ·* ◯ *f*_1_ from the pretrained DCNN model and then form and finetune the new predictor *h*_*t*_ ◯ *f*_*k*_ ◯ *· · ·* ◯ *f*_1_. Our TruncatedTL (TTL) method is illustrated in Fig. 3(i).

The only crucial hyperparameter for TTL is the cutoff index *k*, which depends on the DCNN model and problem under consideration. For SOTA ResNet and DenseNet models that are popularly used in TL for MIC, there are always 4 convolutional blocks each consisting of repeated basic convolutional structures; see, e.g., the bottom of Fig. 3 (ii) for an illustration of ResNet50. So we propose a hierarchical search strategy: **(1) Stage 1: coarse block search**. Take the block cutoffs as candidate cutoff points, and report the best-performing one; **(2) Stage 2: fine-grained layer search**. Search over the neighboring layers of the cutoff from **Stage 1** to optimize the performance. Since *k* is also a crucial algorithm hyperparameter for TF and LWFT, we also adopt the same hierarchical search strategy for them when comparing the performance.

To quickly confirm the efficiency of TTL, we apply TTL and competing TL methods on an X-ray-based COVID-19 classification task on the BIMCV dataset [40]; more details about the setup can be found in Section 4. We pick the COVID example, as the salient radiological patterns in COVID X-rays such as multifocal and bilateral ground glass opacities and consolidations are low-to mid-level visual features [30] and hence we can easily see the benefit of differential TL methods including our TTL. We measure the classification performance by both AUROC (area under the receiver-operating-characteristic curve) and AUPRC (area under the precision-recall curve), and measure the inference complexity by P (number of parameters in the model, M—millions), MACs (multiply-add operation counts [32]^2^, G— billion), T (wallclock run time in milliseconds; details of our computing environment can be found supplementary material).

Table 1 summarizes the results, and we observe that: (1) differential TL methods (TF, LWFT, and TTL) perform better or at least on par with FTL, and the layer-wise search in the second stage further boosts their performance. Also, TTL is the best-performing TL method with the two-stage hierarchical search; (2) TF and TTL that slim down the model lead to reduced model complexity and hence considerably less run time. TTL is a clear winner in terms of both performance and inference complexity. (3) TTL tends to focus on the foreground lesion area instead of the background told by the Grad-CAM visualization.

**Table 1:** A comparison of different TL strategies on BIMCV COVID-19 diagnosis task. (left) a summary of classification performance, (right) visualizations using Grad-CAM.

| Method | AUROC↑ | AUPRC↑ | P(M)↓ | MACs(G)↓ | T(ms)↓ |
| --- | --- | --- | --- | --- | --- |
| FTL | 84.9 ± 0.1 | 85.7 ± 0.3 | 23.5 | 4.12 | 3.59 |
| TF-1 | 85.6 ± 1.1 | 0.863 ± 1.2 | 12.9 | 3.56 | 3.55 |
| LWFT-1 | 848 ± 0.002 | 0.861 ± 0.004 | 23.5 | 4.12 | 3.59 |
| **TTL-1** | **0.851 ± 0.002** | **0.860 ± 0.002** | **8.55** | **3.31** | **3.19** |
| TF-2 | 0.856 ± 0.011 | 0.863 ± 0.012 | 12.9 | 3.56 | 3.56 |
| LWFT-2 | 0.853 ± 0.005 | 0.861 ± 0.001 | 23.5 | 4.12 | 3.56 |
| **TTL-2** | **0.861 ± 0.013** | **0.871 ± 0.008** | **6.31** | **2.87** | **2.97** |

### 3.3 Transferablity analysis

Besides the positive confirmation above, in this section, we provide quantitative corroboration for our claim that top layers might not be needed in TL for MIC. To this end, we need a variant of the classical canonical correlation analysis (CCA), singular-vector CCA (SVCCA) [24].

### SVCCA for quantifying feature correlations

CCA is a classical statistical tool for measuring the linear correlation between random vectors. Suppose that ***x*** *∈* ℝ^*p*^ and ***y*** *∈* ℝ^*q*^ are two random vectors containing *p* and *q* features, respectively. CCA seeks the linear combinations ***u***^⊺^ ***x*** and ***v***^⊺^ ***y*** of the two sets of features with the largest covariance cov(***u***^⊺^ ***x, v***^⊺^ ***y***). Assume 𝔼[***x***] = **0** and 𝔼[***y***] = 0. The problem can be formulated as

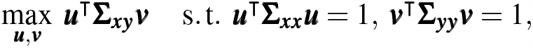

Where 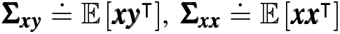, and 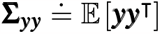,^3^ and the constraints fix the scales of ***u*** and ***v*** so that the objective does not blow up. Once the (***u, v***) pair with the largest covariance is computed, subsequent pairs are computed iteratively in a similar fashion with the additional constraint that new linear combinations are statistically decorrelated with the ones already computed. The overall iterative process can be written compactly as

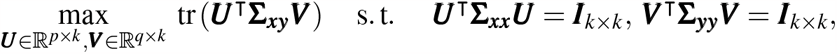

which computes the first *k* pairs of most correlated linear combinations. In practice, all the covariance matrices **Σ**_***xy***_, **Σ**_***xx***_, and **Σ**_***yy***_ are replaced by their finite-sample approximations, and the covariances of the top *k* most correlated pairs are the top *k* singular values of 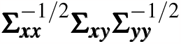 [37], all of which lie in [0, 1]. We call these singular values the *CCA coefficients*. Obviously, high values in these coefficients indicate high levels of correlation.

For our subsequent analyses, we typically need to find the correlation between the features of two data matrices ***X*** *∈* ℝ^*n×p*^ and ***Y*** *∈* ℝ^*n×q*^, where *n* is the number of data points. SVCCA performs principal component analysis (PCA) separately on ***X*** and ***Y*** first, so that potential noise in the data is suppressed and the ensuing CCA analysis becomes more robust. We typically plot the CCA coefficients in descending order when analyzing two group of features.

### Layer transferability analysis

With SVCCA, we are now ready to present quantitative results to show that reusing and finetuning top layers may be unnecessary. We again take the BIMCV dataset for illustration.

#### Features in top layers change substantially, but the changes do not help improve the performance

In Fig. 4 (i) we present the per-layer correlation of the learnt features before and after FTL. First, the correlation monotonically decreases from bottom to top layers, suggesting increasingly dramatic feature finetuning/learning. For features residing in block1 through block3, the correlation levels are substantially higher than that of random features, suggesting considerable feature reuse together with the finetuning. However, for features in block4 which contain the top layers, the correlation level approaches that of random features. So these high-level features are drastically changed during FTL and there is little reuse. The changes do not help and in fact hurt the performance: when we take intermediate features after the FTL and train a classifier based on each level of them, we find that the performance peaks at block3, and starts to degrade afterward. From Fig. 4 (i) (only per-block feature correlations are computed to save space), we find similar patterns in TF, LWFT, and our TTL also: features in bottom blocks are substantially more correlated than those in top blocks, and features in block4—which we remove in our TTL—are almost re-learned as their correlation with the original features come close to that between random features.

**Figure 4.**
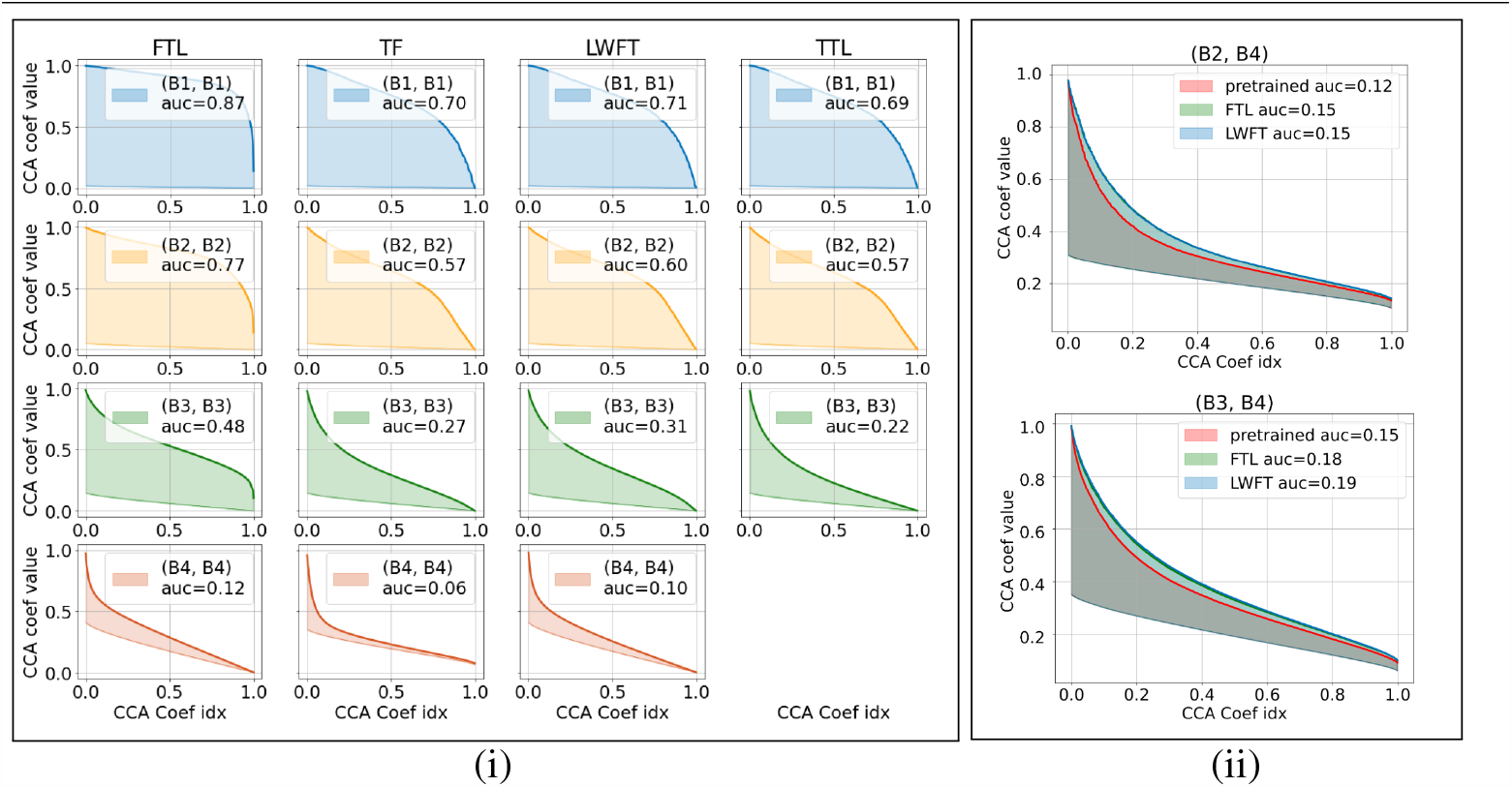
SVCCA on COVID-19 diagnosis task. Bold curve indicates the CCA coefficients for learned features, while light curve indicates the correlation for two uncorrelated random features. So the area between the two curves is a quantitative measure of the correlation between the said blocks. We normalize all the indices of the CCA coefficients to be [0, 1]. We compare (i) features learned from trained and finetuned model at the same layer, and (ii) features learned in the finetuned model but different blocks.

#### Features in top layers become more correlated with bottom layers after TL

The “horizontal” analysis above says the features of the top layers are almost re-learned in FTL, LWFT, and TF, but it remains unclear what features are learned there. If we believe that high-level features are probably not useful for COVID classification [30], a reasonable hypothesis is that these top layers actually learn features that are more correlated with those of lower layers after TL. This seems indeed the case, as shown in Fig. 4 (ii): the correlation level between the block2 and block 4 features, as well as between block3 and block4 features, increases both visibly and quantitatively.

Given the above two sets of findings, our idea in TTL to remove the redundant top layers and keep the essential bottom layers is reasonable toward effective and compact models.

### 4 Experiments

### Experiment setup

We systematically compare FTL, TF, LWFT, and our TTL on 3 MIC tasks covering both 2D and 3D image modalities, and also explore a 2D lung segmentation task. For the 3D MIC task, we choose ResNeXt3D-101 as the default model and compare it with PENet which is a handcrafted model in Huang et al. [12]. Both models are first pretrained on the kinetics-600 dataset [5], and then finetuned with the same setting as in Huang et al. [12]: 0.01 initial LR for randomly initialized weights and 0.1 for pretrained weights, SGD (momentum = 0.9) optimizer, cosine annealing LR scheduler, 100 epochs of training, and best model selected based on the validation AUROC. For experiments involving randomness, we repeat them 3 times and report the mean and standard deviation. More detailed experiment setup, ablation studies, and other explorations can be found in supplementary materials.

### COVID-19 chest x-ray image classification

We take the chest x-rays from the BIMCV-COVID19+ (containing COVID positives) and BIMCV-COVID19*−* (containing COVID negatives) datasets (iteration 1) [40]^4^. We manually remove the small number of lateral views and outliers, leaving 2261 positives and 2463 negatives for our experiment. We have demonstrated the plausibility and superiority of TTL based on this MIC task around Section 3.2, which we do not repeat here.

### Mitotic cells classification

The density of mitotic cells undergoing division (i.e., mitotic figures) is known to be related to tumor proliferation and can be used for tumor prognosis [2]. Since cell division changes its morphology, we expect mid-level blob-like features to be the determinant here. We take the dataset from the mitotic domain generalization challenge (MIDOG2022) [2] which is about detection of mitotic figures, i.e., the training set consists of properly cropped 9501 mitotic figures and 11051 non-mitotic figures, and the task is to localize mitotic figures on large pathological slices during the test. We modify the task into a binary MIC (positives are mitotic figures, and negatives are non-mitotic figures).

Table 2 (top row) summarizes the results. We observe that: **(1)** Our TTL-2 and TTL-1 beat all other TL methods, and also yield the most compact and inference-efficient models; **(2)** Both TTL-1/2 and TF find the best cutoffs at the transition of block3 and block4, implying that high-level features are possibly unnecessary and can even be hurtful for this task and confirming our tuition that mid-level visual features are likely be crucial for decision; **(3)** Of all methods, LWFT with block-wise search performs the worst; even after layer-wise search, its AUROC only matches that of the baseline FTL. The inferior performance of LWFT implies that only fine-tuning the top layers is not sufficient for this task.

**Table 2:** mitotic cell classification with its Grad-CAM visualization (top row), pulmonary embolism CT image classification(bottom left), lung segmentation(bottom right). The best result of each column is colored in **red**. *↑* indicates larger value is better and *↓* indicates lower value is better. “-1” means with the block-wise search only, “-2” means with the two-stage block-layer hierarchical search, “-Bx” means model truncate at block x.

| Method | AUROC↑ | AUPRC↑ | P(M)↓ | MACs(G)↓ | T(ms)↓ |
| --- | --- | --- | --- | --- | --- |
| FTL | 0.925 ± 0.003 | 0.917 ± 0.003 | 23.5 | 4.12 | 3.59 |
| TF-1 | 0.925 ± 0.002 | 0.918 ± 0.002 | 12.9 | 3.56 | 3.50 |
| LWFT-1 | 0.920 ± 0.003 | 0.913 ± 0.005 | 23.5 | 4.12 | 3.55 |
| **TTL-1** | **0.928 ± 0.004** | **0.921 ± 0.003** | **8.55** | **3.31** | **2.99** |
| TF-2 | 0.925 ± 0.002 | 0.918 ± 0.002 | 12.9 | 3.56 | 3.50 |
| LWFT-2 | 0.925 ± 0.001 | 0.919 ± 0.001 | 23.5 | 4.12 | 3.56 |
| **TTL-2** | **0.928 ± 0.004** | **0.921 ± 0.003** | **8.55** | **3.31** | **2.99** |

| Method | AUROC↑ | AUPRC↑ | P(M)↓ | MACs(G)↓ | T(s)↓ |
| --- | --- | --- | --- | --- | --- |
| PENet | 0.822 ± 0.010 | 0.855 ± 0.007 | 28.4 | **51.7** | **1.59e-2** |
| FTL | 0.821 ± 0.010 | 0.867 ± 0.006 | 47.5 | 66.3 | 1.96e-2 |
| TF-1 | 0.849 ± 0.020 | 0.886 ± 0.017 | 36.1 | 64.9 | 1.93e-2 |
| LWFT-1 | 0.817 ± 0.005 | 0.855 ± 0.003 | 47.5 | 66.3 | 1.96e-2 |
| **TTL-1** | **0.854 ± 0.013** | **0.889 ± 0.015** | **26.11** | 60.17 | 1.68e-2 |
| TF-2 | 0.849 ± 0.020 | 0.886 ± 0.017 | 36.1 | 64.9 | 1.93e-2 |
| LWFT-2 | 0.835 ± 0.038 | 0.870 ± 0.028 | 47.5 | 66.3 | 1.96e-2 |
| **TTL-2** | **0.854 ± 0.013** | **0.889 ± 0.015** | **26.11** | 60.17 | 1.68e-2 |

Table 2: mitotic cell classification with its Grad-CAM visualization (top row), pulmonary embolism CT image classification(bottom left), lung segmentation(bottom right).
| Method | Dice Coef ↑ | Jaccard index ↑ | P(M) ↓ | MACs(G) ↓ | T(ms) ↓ |
| --- | --- | --- | --- | --- | --- |
| FTL | 0.968 ± 0.029 | 0.940 ± 0.051 | 24.4 | 5.93 | 11.8 |
| **TTL-B1** | 0.970 ± 0.029 | 0.941 ± 0.052 | **21.3** | **1.68** | **7.05** |
| **TTL-B2** | **0.972 ± 0.027** | **0.946 ± 0.047** | 21.5 | 3.02 | 8.50 |
| **TTL-B3** | 0.968 ± 0.029 | 0.939 ± 0.051 | 22.1 | 4.82 | 10.6 |

### Pulmonary embolism CT image classification

Pulmonary Embolism (PE) is a blockage of the blood vessels connecting the lungs and the heart, and CT pulmonary angiography (CTPA) is the gold standard for its diagnosis [42]. We take the public PE dataset [12] consisting of 1797 CT images from 1773 patients, and compare the performance with handcrafted 3DCNN model, PENet which is the SOTA model that outperforms other 3D DCNN models, such as ResNet3D-50, ResNeXt3D-101, and DenseNet3D-121 as demonstrated in Huang et al. [12].

On CT images, PE often appears as localized blobs that map to mid-level visual features. So the suboptimal TL performance of the SOTA models is mostly likely due to the rigid FTL strategy. We confirm this on ResNeXt3D-101 pretrained on kinetics-600: after layer-wise search, all differential TL methods including TF, LWFT, and TTL outperform PENet by considerable margins in both AUROC and AUPRC, as shown in Table 2 (bottom left). Also, both TTL-1 and TF-1 find the best cutoff at the transition of block3 and block4, another confirmation of our intuition that probably only low-to mid-level features are needed here. We note that although the AUROC obtained via FTL is slightly lower than that of PENet, the AUPRC is actually higher—which [12] does not consider when drawing their conclusion. Our TTL-2 is a clear winner in performance, despite that PENet has been meticulously designed and optimized for the task.

### Chest X-ray lung segmentation

Despite our focus on MIC, we briefly explore the potential of TTL for segmentation also. To this end, we explore a public chest x-ray lung segmentation datasets collected from two source: Montgomery Country XCR set (MC) and Shenzhen Hospital CXR Set (SH) [4, 15], both of which provide manual segmentation masks. MC consists of 58/80 tuberculosis/normal cases, and SH has 336/326 tuberculosis/normal cases.

For simplicity, we only perform block-wise truncation, and our experimental results are shown in Table 2 (bottom right). TTL achieves the best segmentation performance at block 2 (TTL-B2) and outperforms FTL by 0.6% in terms of Dice Coefficient and Jaccard index (both are standard metrics for evaluating segmentation performance). Notably, TTL-B2 is more efficient than FTL and reduces the model size by 12% and inference time by 28%.

## 5 Conclusion

In this paper, we present a thorough examination of transfer learning (TL) in the context of medical image classification (MIC). We introduce a novel method, named TruncatedTL (TTL), which leverages the pre-trained model’s visual semantic representations and aligns them with target medical images. Through a comprehensive analysis of feature transferability, we reveal that in many MIC scenarios, low-to mid-level features suffice for effective transfer learning. We showcase four real-world medical applications, including 2D and 3D MIC (plus one 2D segmentation for exploration), to demonstrate the effectiveness of TTL. Our findings confirm that TTL significantly reduces model complexity, both in terms of size and inference speed, while achieving comparable performance to existing methods. This study contributes to the advancement of transfer learning techniques in the domain of medical imaging and underscores the potential of TTL as a promising alternative for improving model performance and efficiency in MIC tasks.

## Supporting information

supplemental results

## Data Availability

https://bimcv.cipf.es/bimcv-projects/bimcv-covid19/
https://stanfordmlgroup.github.io/competitions/chexpert/
https://www.kaggle.com/datasets/yoctoman/shcxr-lung-mask
https://academictorrents.com/details/ac786f74878a5775c81d490b23842fd4736bfe33
https://imig.science/midog/

https://stanfordmlgroup.github.io/competitions/chexpert/

https://www.kaggle.com/datasets/yoctoman/shcxr-lung-mask

https://academictorrents.com/details/ac786f74878a5775c81d490b23842fd4736bfe33

https://imig.science/midog/

## Footnotes

1 Finetuning starts with the original pretrained weights each time.

2 https://github.com/sovrasov/flops-counter.pytorch

3 ≐ means “defined as”.

4 https://bimcv.cipf.es/bimcv-projects/bimcv-covid19/#1590858128006-9e640421-671

