## supplemental results for "Rethinking Transfer Learning for Medical Image Classification"

#### A Additional experimental details

##### A.1 Experimental protocol

**Training TL models** All 2D experiments follow the same training protocol unless otherwise stated: **1)** the given dataset is split into 64% training, 16% validation, and 20% test; **2)** ResNet50 is the default model for all 2D MIC tasks, and ResNet34 for 2D segmentation; **3)** all 2D images are center-cropped and resized to $224 \times 224$; **4)** random cropping and slight random rotation are used for data augmentation during TL; **5)** the ADAM optimizer is used for all TL methods, with an initial LR $10^{-4}$ and a batch size 64; The `ReduceLROnPlateau` scheduler in PyTorch is applied to adaptively adjust the LR: when the validation AURPC stagnates, the LR is decreased by $1/2$. Training is terminated when the LR drops below $10^{-7}$; **6)** the best model is chosen based on the validation AUPRC.

**Evaluating TL models** We measure the runtime on a system with Intel Core i9-9920X CPU and Quadro RTX 6000 GPU. We report AUROC/AUPRC for MIC, and Dice Coefficient/Jaccard Index for segmentation as performance metrics and Params, MACs, runtime time as complexity metrics.

##### A.2 Extending TTL on segmentation models

We construct the TTL model in a manner similar to what we have done for the MIC tasks. We take the U-Net architecture with ResNet34 and pretrained on ImageNet as our backbone

© 2023. The copyright of this document resides with its authors.
It may be distributed unchanged freely in print or electronic forms.

model. To ensure the skip-connection structure can be preserved after truncation, we symmetrically truncate the backbone and segmentation head simultaneously. We do not include a comparison with LWFT and TF, as the original papers do not discuss how to extend them to segmentation and our MIC tasks above already show the superiority of our TTL.

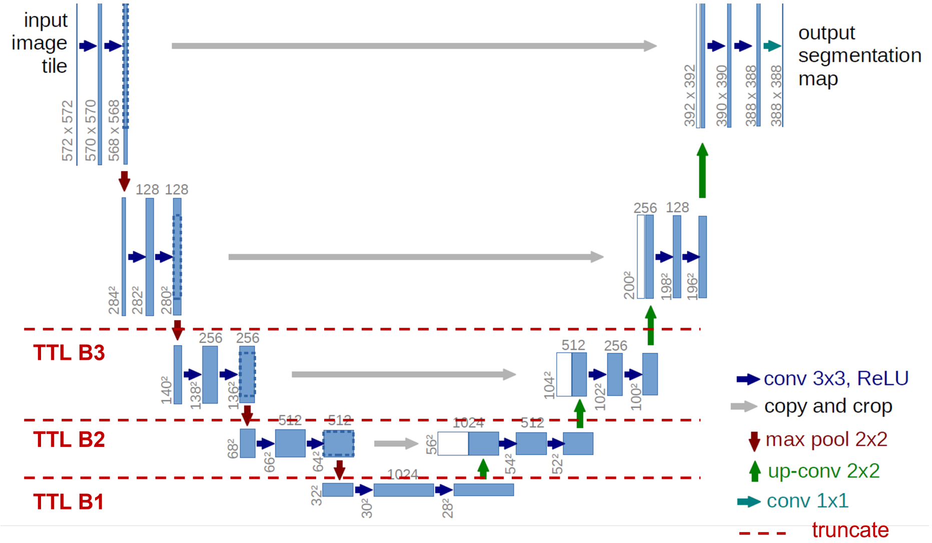


Figure 5: Similar to TTL on ResNet/DenseNet, we identify the block structure in U-Net and truncate at the intersection between blocks. The image of U-Net is adapted from Ronneberger et al. [2]

### B Ablation studies

#### B.1 Impact of network architecture

To study the impact of network architecture on the result of TTL, we compare three network models: ResNet50, DenseNet121, and EfficientNet-b0, which represent large-, mid-, and small-size models, respectively, on COVID-19 classification. The results are summarized in Table 3. It is clear that our TTL uniformly improves the model performance and reduces the model size by at least several fold, compared to FTL. In particular, even for the most lightweight model EfficientNet-b0, TTL still pushes up the performance by about 5% in both AUROC and AUPRC, and reduces the model size by about 4-fold.

#### B.2 Impact of pooling

After truncation, we can either directly pass the full set of features or downsample them before feeding them into the MLP classifier. For deep learning models, we can easily perform subsampling by inserting a pooling layer. For this purpose, we use PyTorch's built-in function `AdaptiveAvgPool2d` to downsample the feature map to $1 \times 1$ in the spatial dimension, which produces the most compact spatial features. We compare models with

Table 3: Impact of network architecture on TTL performance. The best results in each group are colored in **red**.

| | Method | AUROC | AUPRC | Params(M) | MACs(G) | Speed(s) CPU | Speed(s) GPU |
|---|---|---|---|---|---|---|---|
| RN50[1] | FTL | $0.853 \pm 0.004$ | $0.861 \pm 0.002$ | 23.5 | 4.12 | 80.0 | 6.72 |
| | TTL | **$0.865 \pm 0.001$** | **$0.870 \pm 0.007$** | **8.55** | **3.31** | **60.9** | **5.83** |
| DN121[2] | FTL | $0.852 \pm 0.002$ | $0.856 \pm 0.004$ | 18.2 | 3.31 | 107 | 24.0 |
| | TTL | **$0.866 \pm 0.006$** | **$0.871 \pm 0.012$** | **1.53** | **2.11** | **46.2** | **5.94** |
| ENb0[3] | TL | $0.795 \pm 0.004$ | $0.794 \pm 0.002$ | 3.63 | 0.378 | 30.0 | 8.33 |
| | TTL | **$0.842 \pm 0.001$** | **$0.843 \pm 0.007$** | **0.896** | **0.255** | **24.0** | **6.46** |

Abbreviation: [1] ResNet50, [2]:DenseNet121, [3] EfficientNet B0

vs. without this extra pooling layer on COVID-19 classification, and present the results in Fig. 6. We observe no significant gap between the two settings in terms of peak performance measured by both AUPRC and AUROC. However, the setting with pooling induces a lower-dimensional input to the MLP classifier, and hence contains much fewer trainable parameters compared to that without pooling.

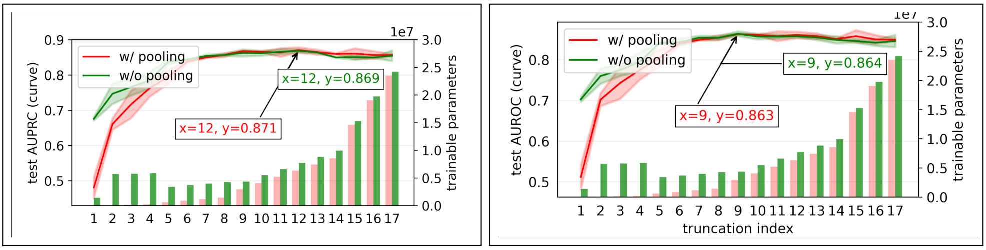


(i) (ii)

Figure 6: ResNet50 transferred from ImageNet to BIMCV using our TTL. We report the performance in both AURPC (i) and AUROC (ii). The red and green curves represent TTL performance with and without the extra adaptive pooling layer between the final convolution layer and the MLP classifier. Arrows point to the peak performance.

### C Exploratory studies

#### C.1 Detecting the near-best truncation point

Our two-stage search strategy for good truncation points (see Section 3.2) is effective as verified above, but can be costly due to the need for multiple rounds of training, each for one candidate truncation point. In this subsection, we explore an alternative strategy to cut down computation: the idea is to detect the transition point where feature reuse becomes negligible. For this, we recall the SVCCA analysis in Fig. 4: we use the distribution of the SVCCA coefficients to quantify the correlation of features before and after FTL. To determine when the correlation becomes sufficiently small, we compare the correlation with that between random features.

We quickly test the new strategy on both COVID-19 and CT-based PE classification, shown in Fig. 7 left and Fig. 7 right, respectively. From Fig. 7, we can see that for COVID-19 classification, feature reuse diminished after the 12-th candidate truncation point: from

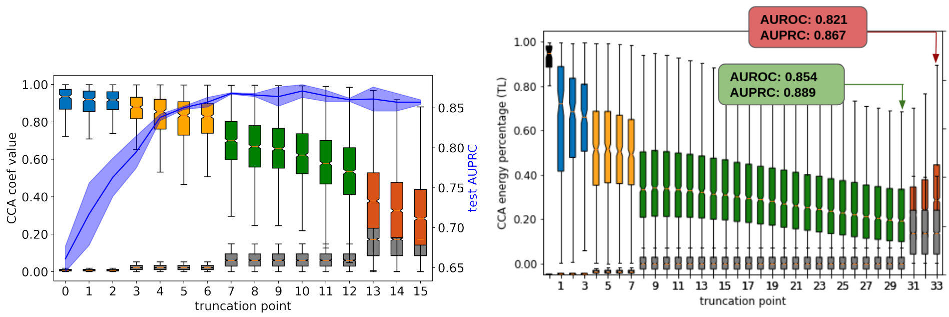


Figure 7: **Left**: SVCCA on ResNet50 transferred from ImageNet to BIMCV. The plot is inherited from Fig. 3 (ii). The gray boxes and the associated bars represent the distributions of CCA coefficients between random features. **Right**: SVCCA on ResNeXt3D-101 transferred from Knetic-600 to CTPA. Truncating at layer 30 (marked in green) significantly outperforms FTL (marked in red).

13-th onward, feature correlation becomes very close to that between random features, suggesting that the features are almost uncorrelated. As expected, the 12-th truncation point also achieves near-optimal performance as measured by AUPRC. Similarly, for CT-based PE classification in Fig. 7 right, this strategy suggests the 30-th truncation point, which yields the same result as that we have obtained using the two-stage search strategy as reported in Table 2 bottom left, in both AUPRC and AUROC. Note that in Fig. 7 right, the SVCCA coefficients slightly increase after the 30-th truncation point due to dimensionality: the channel sizes are doubled there compared to the previous blocks, and the raised dimension induces higher correlation coefficients. But to decide if feature reuse becomes trivial, we only need to check if the two SVCCA-coefficient distributions are sufficiently close, not their absolute scalings.

Thus, this new strategy seems promising as a low-cost alternative to our default two-stage search strategy. We summarize the key steps as follows.

**Step 1**: perform FTL on the target dataset;

**Step 2**: compute SVCCA coefficients between the features before and after FTL, at all candidate truncation points;

**Step 3**: compute SVCCA coefficients between random features, at all candidate truncation points;

**Step 4**: locate the truncation point where the distribution of the two groups of SVCCA coefficients from **Step 2** and **Step 3** becomes substantially overlapped, and take it as the truncation point;

**Step 5**: fine-tune the model with the truncation point selected from **Step 4**.

Compared to our two-stage strategy that entails numerous rounds of model finetuning, the new strategy only requires two rounds of finetuning: one from **Step 1** on the whole model, and the other from **Step 5** on the truncated model.

#### C.2 Inter-domain TL

All we have discussed above are intra-domain TL where the source domain is distinct from the target domain. It is tempting to think inter-domain TL might be more effective. To

Table 4: Symptom of lung disease in CheXpert

| Diseases | Shape | Edge | Contrast | level of features |
|---|---|---|---|---|
| No Finding | ☒ | ☒ | ☒ | None |
| Enlarged Cardiom. | ☑ | ☒ | ☑ | low and high |
| Cardiomegaly | ☑ | ☒ | ☑ | low and high |
| Lung Opacity | ☒ | ☒ | ☑ | low |
| Lung Lesion | ☑ | ☒ | ☑ | low and high |
| Edema | ☒ | ☒ | ☑ | low and high |
| Consolidation | ☒ | ☒ | ☑ | low |
| Pneumonia | ☒ | ☒ | ☑ | low |
| Atelectasis | ☑ | ☒ | ☑ | low and high |
| Pneumonthorax | ☒ | ☑ | ☑ | low |
| Pleural Effusion | ☒ | ☒ | ☑ | low |
| Pleural Other | ☒ | ☒ | ☑ | low |
| Fracture | ☑ | ☑ | ☑ | low and high |
| Support device | ☑ | ☑ | ☑ | low and high |

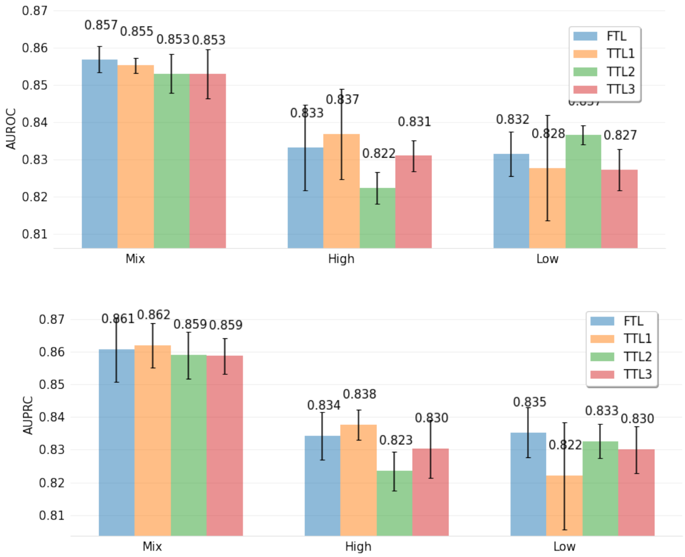


Figure 8: ResNet50 transferred from CheXpert to BIMCV. (top) test AUROC score; (bottom) test AUPRC score. “Mix", “High", and “Low" mean finetuning from pretrained models on the original CheXpert, CheXpert-high, and CheXpert-low, respectively. TTLx means the truncation point is chosen at the transition point between block x and block x+1.

test this, we take the x-ray-based COVID-19 classification task again, but now finetune the ResNet50 model pretrained on CheXpert, a massive-scale x-ray dataset ($\sim 220K$ images) for disease prediction covering 13 types of diseases (see Table 4) [1]. These diseases correspond to varying levels of visual features. Hence, we categorize them into two groups: **CheXpert-low** includes diseases that need low-level features only, and **CheXpert-high** covers those needing both low- and high-level features. A summary of the categorization can be found in Table 4[1]. We pretrain ResNet50 on 3 variants of the dataset, respectively: full CheXpert, CheXpert-low, and CheXpert-high, and then compare FTL and our TTL on the three resulting models. For TTL, we only perform a coarse-scale search and pick the 3 transition layers between the 4 blocks in ResNet50 as truncation points. As always, all experiments are repeated three times.

From the results summarized in Fig. 8, we find that: **(1)** Our TTL always achieves comparable or superior performance compared to FTL, reaffirming our conclusion above; **(2)** Transferring from the model pretrained on the original dataset substantially outperforms transferring from those pretrained on the CheXpert-high and CheXpert-low subsets. This can be explained by the diversity of feature levels learned during pretraining: on the subsets more specialized features are learned; in particular, on CheXpert-low, perhaps only relatively low-level features are learned; and **(3)** Notably, our results show that medical-to-medical TL does not do better than TL from natural images, when we compare the results in Fig. 8 with those in Table 1. We suspect this is because feature diversity is the most crucial quality required on the pretrained models in TL, and models pretrained on natural images perhaps already learn sufficiently diverse visual features.

### D Sample images of Grad-CAM analysis

To find out what visual features are learned by different models, we conduct a feature analysis study using Grad-CAM[3]. Our empirical findings show that deep models tend to produce spatially extensive features that spread over the image. In contrast, shallow models such as those produced by TTL tend to find spatially localized features that often capture the infected area around the lungs and the mitotic region, as shown in Fig. 1 and Table 2. In particular, those localized regions often take the form of the abnormal texture of boundary changes and are invariant to the global object (e.g., the shape of the ribs and cells), which demonstrates the effectiveness of TTL in learning more discriminative features for MIC.

### E Feature visualization on pretrained CNN

To gain insights into the features acquired by layers in a CNN pre-trained on a general natural image dataset, we employed visualization techniques to elucidate the learned features across various layers in a VGG19 network pretrained on ImageNet. The results are shown in Fig. 9. Our empirical observation reveals that bottom layers (e.g., layer 1, 5, and 10) mostly learn low-semantic patterns such as color and simple structured patterns. As delve deeper into top layers (e.g., layer 20 and 30), it progressively refines its knowledge, capturing higher-level semantic characteristics, including specific object-related local regions.

[1]Disease symptoms information obtained from Mayo clinic: https://www.mayoclinic.org/symptom-checker/select-symptom/itt-20009075

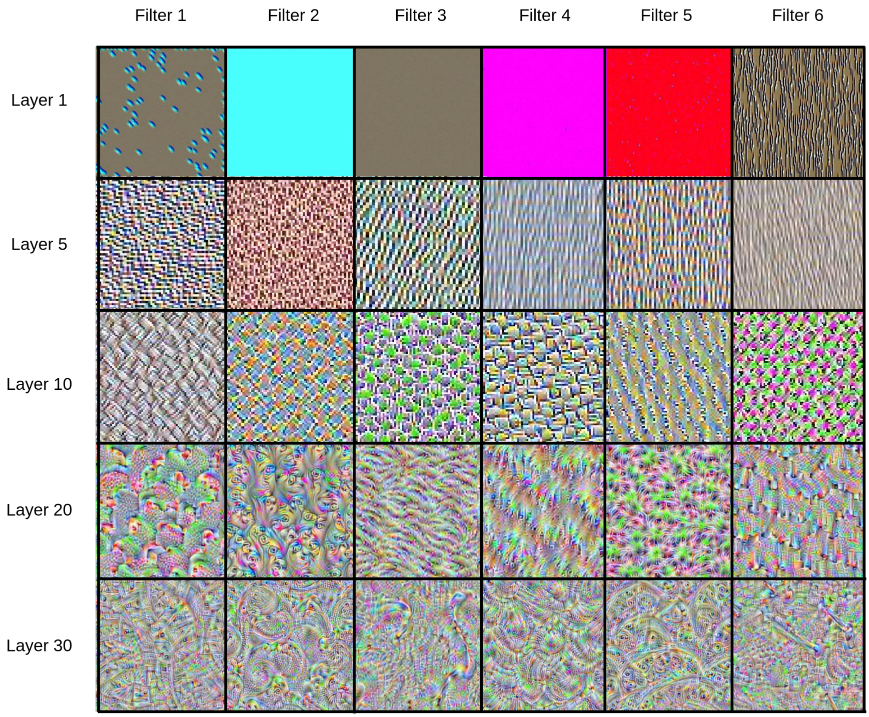


Figure 9: Visualized features learnt at different layers in VGG19 pretrained on ImageNet. To get the visualized images, we optimize the inputs with respect to the output value from a certain filter.
